# Comparing angiogenic biomarkers and clinical history of hypertensive disease of pregnancy with the association of hypertension up to 15 years after delivery

**DOI:** 10.1101/2025.07.22.25330603

**Authors:** Tooba Anwer, David E. Cantonwine, Ellen W. Seely, Kathryn Gray, Thomas F. McElrath

## Abstract

**Objective:** We investigated the independent and comparative association of a history of hypertensive disease of pregnancy and elevated serum soluble fms-like tyrosine kinase 1 to placental growth factor (sFlt-1/PlGF) ratio during the second half of pregnancy with the development of HTN up to 15 years after delivery.

Study Design: N=1,238 singleton pregnancies were part of a prospective birth cohort study that enrolled patients from 2006-2008. Serum sFlt-1/PlGF was collected at a median of 26.0 and 35.1 weeks gestation. Adjusted Cox proportional hazard models estimated hazard ratios and 95%CI for the time to diagnosis for Stage 1 and Stage 2 HTN within 15 years PP in association with diagnoses of HDP and interquartile range of sFlt-1/PlGF. We adjusted for confounders.

**Results:** Of the n=993 women, n=260 (29.18%) were diagnosed with stage 1 and n=169 (17.0%) with Stage 2 HTN within 15 years of delivering.A history of PE or a history of gestational hypertension were both significantly associated with developing HTN later in life with adjusted hazard ratios of 2.17 (95% CI, 1.44-3.28) and 3.50 (95% CI, 2.21-5.54). There was no significant association between the hazard of developing HTN and the sFlt/PlGF ratio. However, mean PlGF levels during pregnancy in those who remained normotensive in the follow-up were higher at 546.7 pg/mL (SD= 369.5) compared to 513.7 pg/mL (SD=389.9) in the group with stage 1 and stage 2 HTN combined and 471.7 pg/mL (SD=471.7) in the stage 2 only group (p=0.008 and p=0.001).

**Conclusion:** In patients followed for up to 15 years after delivery, a clinical history of hypertensive disease of pregnancy was significantly associated with the hazard of developing HTN later in life, while elevated sFlt-1/PlGF biomarker levels were not.However, mean PlGF levels in the latter half of pregnancy were significantly lower in those who developed HTN later in life.

## INTRODUCTION

Hypertensive disorders of pregnancy (HDP) have a global prevalence of 116 per 100,000 women of childbearing age^1^ and are the leading cause of maternal morbidity and mortality worldwide^2^. HDP definition includes a diagnosis of chronic hypertension (HTN), white coat HTN, masked HTN, gestational hypertension (GHTN), and preeclampsia (PE) according to the International Society for the Study of Hypertension in Pregnancy^3^. The association between HDP and long terms risks of cardiovascular disease (CVD) is well established. PE is associated with a two-fold rise in risk of coronary heart disease, stroke, and death from CVD and four-fold increase in future heart failure.^4,5^ Specifically, pregnancies complicated by PE increase the risk of incident CVD within 9 years of delivery, left ventricular hypertrophy, and higher blood pressure (BP) postpartum (PP)^6– 8^. A study found a fivefold increase in HTN rates within five years of a pregnancy complicated by PE^9^, and another found that up to one-third of women with PE may develop HTN within a decade of an affected pregnancy^10^ Gestational hypertension (GHTN) is also associated with a 1.8 fold increase in risk of coronary heart disease and heart failure and a 1.4 fold increase in composite CVD^11,12^. Given the nearly doubled incidence of PE and GHTN in the US from 2007 to 2019^13,14^, predicting when long-term adverse cardiovascular events occur later in life is crucial.

Several studies suggest that angiogenic factor imbalance is involved in the pathophysiology of PE^15–19^. The PRAECIS study found that, in pregnant women with hypertensive disease of pregnancy between 23 and 35 weeks of gestation, measurement of a ratio of serum soluble fms-like tyrosine kinase-1 to placental growth factor (sFlt-1/PlGF) provided stratification of the risk of progressing to severe PE within the coming fortnight along with a strong association with adverse outcomes^20^. sFlt-1/PlGF testing during pregnancy has emerged as a pivotal biomarker for predicting severe PE within two weeks, leading to the development of a commercial test. However, the extent to which angiogenic imbalance during pregnancy correlates with the future risk of CVD, particularly HTN, remains an area of ongoing research. A small prospective study of 40 women with a history of PE, found that elevated serum sFlt-1/PlGF at the time of diagnosis was correlated with the mean diastolic blood pressure 12 years PP^21^.

Both the American Heart Association (AHA) and the European Society of Cardiology (ESC) recognize that a clinical history of HDP increases the risk of CVD. However, it is uncertain how objective patient clinical data in pregnancy can be used to understand the risk of HTN and later CVD. Screening and stratifying this risk with biomarkers collected during pregnancy may guide patients towards appropriate care after delivery. This study aimed to determine if a history of HDP or angiogenic biomarkers, measured during pregnancy and irrespective of diagnosis of HDP, was associated with the development of HTN up to 15 years after delivery. We hypothesized that angiogenic biomarkers, specifically elevated serum sFlt-1/PlGF levels, measured during the second and third trimester of pregnancy would add to prediction of subsequent HTN compared to a history of HDP alone.

## METHODS

### Study Design

We performed a retrospective cohort study selected from the ongoing LIFECODES pregnancy biobank at Brigham and Women’s Hospital (BWH), Boston, MA. Methods pertaining to patient recruitment, sample collection and clinical data curation for LIFECODES are detailed in prior publications^22,23^. Briefly, participants were eligible for recruitment at their initial prenatal visit if they were ≥18 years of age, had no higher order pregnancy greater than twins, and were receiving prenatal care and planned to deliver at BWH. This retrospective cohort study included those enrolled between 2006-2008 who delivered a live born infant and did not have pre-existing HTN (N=1,238). N=245 subjects were not included in the analyses due to missing sFlt-1/PlGF values (N=99) in the database or missing BP measurements postpartum (N=146) in the electronic medical records (EMR). N=993 subjects were included in the final analyses. A diagnosis of HDP (consisting of GHTN and PE) developed in the index pregnancy was also abstracted from the database. This study was approved by the institutional review board at the Mass General Brigham HealthCare System, and written informed consent was obtained from all participants (2009P000810).

### Assessment of Angiogenic Biomarkers

Concentrations of circulating PlGF and sFlt-1 were measured in maternal EDTA plasma samples collected in the second and third trimester. The median gestational week of sampling was 26.0 (25.0–26.9) and 35.1 (34.4–35.9) weeks. PlGF and sFlt-1 levels were measured with prototype ARCHITECT immunoassays (Abbott Laboratories) ^24^.The PlGF immunoassay measures unbound PlGF (i.e., free form of PlGF-1), with a lower limit of detection (LLOD) of 1 pg/mL and an upper limit of quantification of (ULOQ) of 1,500 pg/mL. The sFlt-1 immunoassay measures total sFlt-1 (i.e., both free and bound sFlt-1), with a LLOD of 0.10 ng/mL and an ULOQ of 150 ng/mL^25^.

### Covariates

Self-reported questionnaires were administered at enrollment, which included sociodemographic factors (e.g., maternal age, race, ethnicity, and maternal education) and behavioral factors (e.g., smoking and alcohol use during pregnancy). Past medical history, vitals, and labor and delivery information were abstracted from the EMR. Self-reported race included the following categories: White, Black, South Asian, East Asian, Native American/Pacific Islander, more than one race, other, unsure, and N/A. Hispanic ethnicity was recorded separately. In our analysis, we categorized race and ethnicity as non-Hispanic White, non-Hispanic Black, Hispanic, Asian, and other. Pregnancy dating was confirmed by ultrasound performed less than or equal to 12 weeks gestation (GA). If consistent with the last menstrual period (LMP) dating, the LMP was used to determine the due date. If not consistent, then the due date was set by the earliest available ultrasound ≤ 12 weeks gestation. Past medical history and relevant pregnancy conditions were independently reviewed by two Maternal–Fetal Medicine faculty physicians. Potential confounders were selected based on directed acyclic graphs (DAGs) presented in supplement 1: the minimally sufficient adjustment set included age at enrollment (years), pre-pregnancy BMI (kg/m^2^), self-identified race and ethnicity, insurance status (private vs. public/none), systolic blood pressure at enrollment (mmHg), smoking during pregnancy (yes/no), and diagnosis of PE or GHTN in current pregnancy (yes/no).

### Definition of Hypertension

The diagnostic criteria for HTN as defined by the ACC and AHA were used as outlined in Table 1^26^. We examined both stage 1 and stage 2 HTN given previous studies have used the older definition of HTN, defined as systolic blood pressure (SBP) ≥ 140 mmHg or diastolic blood pressure (DBP) ≥ 90 mmHg. A patient was given a diagnosis of HTN if: 1) EMR review revealed a diagnosis of HTN listed in the problem list or visit notes, or 2) there were 2 or more hypertensive BP measurements documented in vitals, or 3) had an anti-hypertensive medication in their medication list, which was then confirmed in the chart to be prescribed for HTN. The BP readings for each subject were collected through a comprehensive chart review of the electronic medical record (EMR). The review period for each patient spanned from six weeks postpartum from the index pregnancy to December 2022. The diagnosis of HTN was established if a participant had a BP with a systolic BP ≥ 130 mm Hg or a diastolic BP ≥ 80 mm Hg documented on at least two separate occasions in the electronic medical record ≥ 6 weeks postpartum. BPs collected in subsequent pregnancies and < 6 weeks postpartum were excluded when establishing the diagnosis of HTN. Furthermore, the diagnosis of HTN was further stratified into stage 1 and stage 2 HTN according to the ACC/AHA criteria. The diagnosis of HTN was individually confirmed for each participant by a reviewer.

**Table 1.** Classification of Hypertension

| Classification of Hypertension |  |  |  |
| --- | --- | --- | --- |
| Systolic BP (mmHg) |  | Diastolic BP (mmHg) | ACC/AHA (2017) |
| < 120 | AND | < 80 | Normal |
| 120-129 | OR | <80 | Elevated BP |
| 130-139 | OR | 80-90 | Stage 1 Hypertension |
| ≥ 140 | OR | ≥ 90 | Stage 2 Hypertension |

### Definition of Gestational Hypertension and Preeclampsia

GHTN was defined as systolic BP ≥ 140 mmHg or diastolic BP ≥ 90 mmHg diastolic after 20 weeks of gestation on two separate occasions ≥ 4 hours apart. PE was defined as systolic BP ≥ 140 mmHg or diastolic BP ≥ 90 mmHg after 20 weeks of gestation on two separate occasions ≥ 4 hours apart, along with positive urinary protein testing (> 300 mg/24 hr or a spot protein/creatinine ratio > 0.30)^27^. PE was further categorized into early-onset (diagnosis prior to 34 weeks gestation) vs. late onset (diagnosis ≥ 34 weeks gestation). All cases of PE were de-identified and independently reviewed by two Maternal–Fetal Medicine faculty physicians. In the case of disagreement, a third MFM physician reviewed the case.

### Statistical Analysis

Analyses were performed using SAS version 9.4 (SAS Institute Inc., Cary, NC). *p*-Values < 0.05 were used to define statistical significance. Sociodemographic and clinical characteristics of the participating women were described, and associations between those with follow up measurements and those either excluded or without follow up measurements were examined using chi-square, Fisher’s exact, or Wilcoxon rank-sum test, as appropriate. Values of sFlt-1, PlGF and the ratio of sFlt-1/PlGF between those diagnosed as Stage 1 HTN or Stage 2 HTN were compared to normotensive participants using Wilcoxon rank-sum test. Unadjusted Cox proportional hazard regression models were run with combined and separated stage 1 and 2 HTN as the outcome to calculate hazard ratios corresponding to a diagnosis of early onset PE, late onset PE, all PE, and GHTN. In an adjusted model, maternal age at enrollment, pre-pregnancy BMI, systolic blood pressure at enrollment, and health insurance category were covariates.

Similarly, unadjusted Cox proportional hazard regression models were run with combined and separated stage 1 and 2 HTN as the outcome to calculate hazard ratios (HR) corresponding to an interquartile range (IQR) increase in sFlt-1, PlGF, and/or sFlt-1/PLGF ratio during the second and third trimester. In fully adjusted models, maternal age at enrollment, pre-pregnancy BMI, systolic blood pressure at enrollment, race/ethnicity, health insurance category, diagnosis of PE or GHTN in current pregnancy (yes/no), and smoking status (yes/no) were covariates.

## RESULTS

Of the 1238 individuals aged 18-50 years who were enrolled in LIFECODES from 2006 to 2008, 245 individuals were either lost to follow-up and did not have blood pressure data available in the electronic medical records from 6 weeks postpartum through December 2022 or were missing sFlt-1/PlGF concentrations (Figure 1). Of the 113 individuals with a HDP diagnosis, 8 had early-onset PE, 56 had late-onset PE, and 49 had GHTN. Baseline demographics were compared between the group with follow-up BP data and those missing follow-up BP data (Table 2). The groups were similar in maternal age, race, education history, mode of delivery, neonatal sex, and type of insurance.

**Table 2.** Baseline Characteristics of Participants Included and Excluded from Study

| Participant characteristics | Participants with follow-up (n=993) | Participants excluded/no follow-up (n=245) | p-value<sup>a</sup> |
| --- | --- | --- | --- |
|  | Mean (SD) or N (%) | Mean (SD) or N (%) |  |
| Maternal age (y) | 31.8 (5.7) | 31.6 (6.0) | 0.37 |
| Maternal pre-pregnancy BMI (kg/m<sup>2</sup>) | 25.0 (5.4) | 25.3 (5.5) | 0.51 |
| Maternal race/ethnicity |  |  |  |
| White/non-Hispanic | 624 (62.8) | 151 (61.6) | 0.53 |
| Black/non-Hispanic | 123 (12.4) | 40 (16.3) |  |
| Asian | 68 (6.9) | 17 (6.9) |  |
| Hispanic | 128 (12.9) | 28 (11.4) |  |
| Other/Mixed | 50 (5.0) | 9 (3.7) |  |
| Insurance Status |  |  |  |
| Private/HMO | 757 (79.3) | 187 (78.9) | 0.38 |
| MassHealth/Medicaid/Self-pay | 198 (20.7) | 50 (21.1) |  |
| Maternal education |  |  |  |
| High school or less | 169 (17.0) | 50 (20.4) | 0.28 |
| Some college/technical school | 146 (14.7) | 34 (13.9) |  |
| College or greater | 678 (68.3) | 161 (65.7) |  |
| Use of Assisted Reproductive Technology | 103 (10.4) | 18 (7.4) | 0.15 |
| Nulliparous | 437 (44.0) | 101 (41.2) | 0.43 |
| History of preeclampsia diagnosis | 27 (4.9) | 8 (5.6) | 0.67 |
| Smoked during pregnancy | 50 (5.0) | 14 (5.8) | 0.63 |
| Alcohol consumption during index pregnancy | 38 (3.9) | 6 (2.6) | 0.44 |
| Preeclampsia diagnosis during index pregnancy | 64 (6.5) | 7 (2.9) | 0.03 |
| Gestational hypertension diagnoses during index pregnancy | 49 (4.9) | 1 (0.4) | 0.0004 |
| A study population with follow up versus missing follow up compared utilizing T test for normally distributed data, Wilcoxon rank sum test for non-parametric data, chi squared and Fishers exact for categorical data, or ANOVA where appropriate |  |  |  |

**Figure 1.**
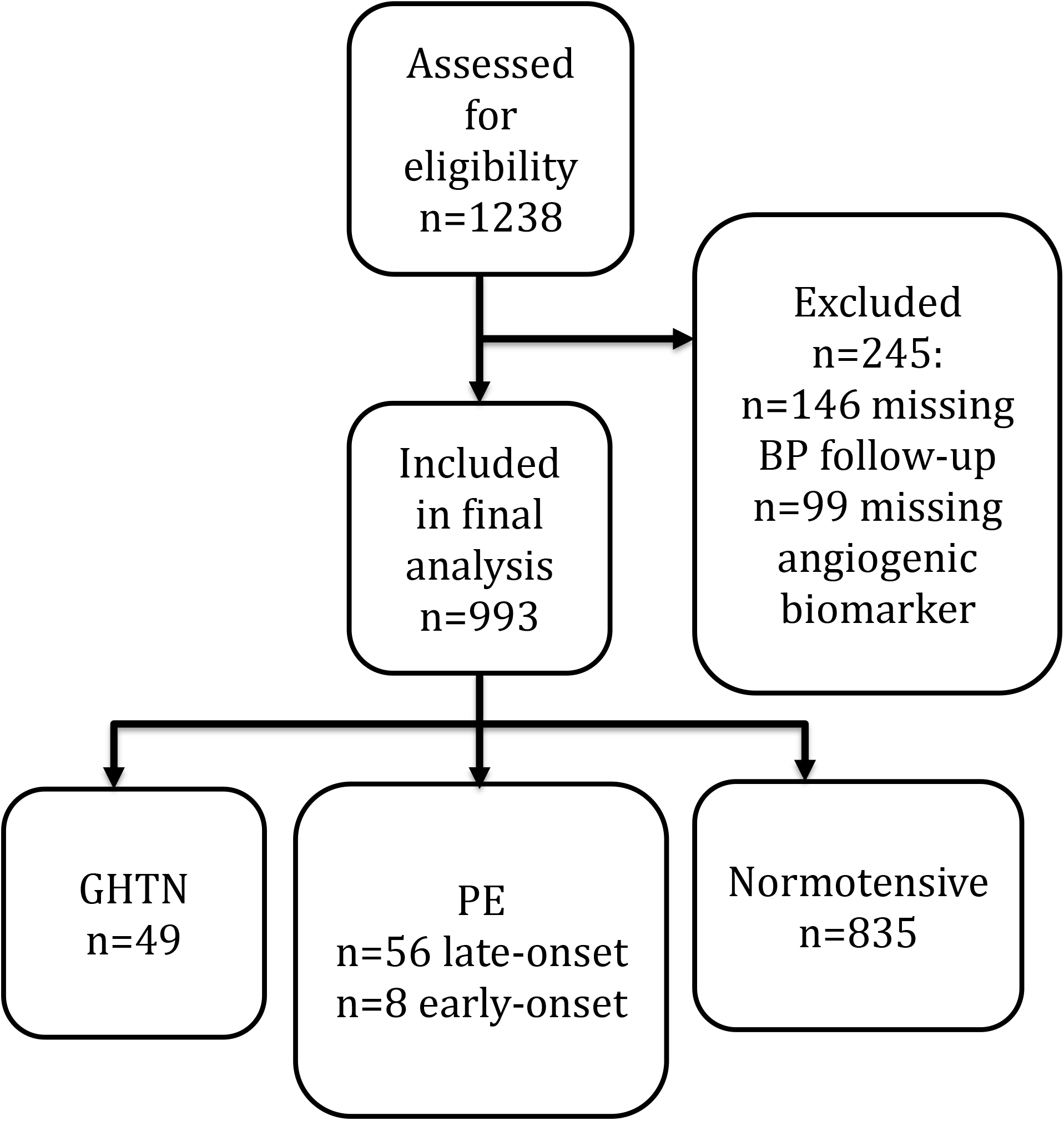
Flow Diagram for Selection of the Cohort of Patients in Study

A total of 993 individuals had serum sFlt-1 levels, PlGF levels, and sFlt-1/PlGF ratios collected at visits throughout pregnancy. Of these n=993 women, n=260 (29.18%) were diagnosed with Stage 1 HTN and n=169 (17.0%) were diagnosed with Stage 2 HTN within 15 years of delivering.

Table 3 demonstrates the hazard of developing HTN in the 15 years after a pregnancy complicated by HDP. PE carries an elevated hazard of developing stage 1 or stage 2 HTN combined later in life, both before and after adjusting for time varying age, BMI at the start of pregnancy, SBP at initial visit, and insurance type, with an adjusted HR of 2.17 (95% CI, 1.44-3.28). This was driven mostly by a diagnosis of late-onset PE. GHTN, however, carried a higher hazard of HTN compared to all PE, with an adjusted of HR 3.50 (95% CI, 2.21-5.54). These trends were similar when looking at the hazard of HTN for the development of stage 2 HTN only.

**Table 3.** Hazard of developing hypertension based on type of hypertensive disease of pregnancy over a 15 year follow-up period

|  | Unadjusted HR (95% CI) | Adjusted HR (95% CI) |
| --- | --- | --- |
| <u>Stage 1 and 2 CHTN</u> |  |  |
| All PE | 3.66 (2.69, 4.98) | 2.17 (1.44, 3.28) |
| Early-onset PE | 8.75 (3.25, 23.55) | 1.80 (0.25, 13.00) |
| Late-onset PE | 3.69 (2.67, 5.09) | 2.41 (1.58, 3.67) |
| GHTN | 2.87 (2.04, 4.04) | 3.50 (2.21, 5.54) |
| <u>Stage 2 only CHTN</u> |  |  |
| All PE | 6.24 (4.29, 9.10) | 3.12 (1.85, 5.26) |
| Early-onset PE | 26.80 (8.49, 84.74) | 9.31 (1.27, 68.30) |
| Late-onset PE | 6.20 (4.18, 9.20) | 3.34 (1.94, 5.75) |
| GHTN | 3.22 (1.96, 5.30) | 5.15 (2.74, 9.68) |
| CHTN= chronic hypertension, PE=preeclampsia, GHTN= gestational hypertension<br><sup>a</sup> Adjusted for time varying age and BMI, systolic blood pressure at initial visit, and insurance. |  |  |

The mean serum sFlt-1 levels, PlGF levels, and ratios are displayed based on timing of collection during the second and third trimester and whether the individuals were normotensive, had stage 2 HTN, or developed any HTN (stage 1 and stage 2) combined during the follow-up period of up to 15 years (Table 4). sFlt-1/PlGF ratios in the second trimester were similar amongst those who remained normotensive (0.02; SD=0.03), developed stage 1 and stage 2 HTN combined (0.02; SD=0.06), and stage 2 HTN only (0.03; SD=0.06) and did not correlate to development of stage 2 HTN alone or stage 1 and stage 2 HTN combined later in life (p=0.062 and p=0.23). This trend was the same for ratios in the third trimester. PlGF levels were significantly higher in the individuals that remained normotensive in the follow-up period compared to those who developed HTN. Specifically, mean PlGF was 546.7 pg/mL (SD= 369.5) in the normotensive group, 513.7 pg/mL (SD=389.9) in the stage 1 and stage 2 HTN combined group, and 471.7 pg/mL (SD=471.7) in the stage 2 only group (p=0.008 and p=0.001). This trend was similar for PlGF concentrations in the third trimester but was not statistically significant.

**Table 4.**
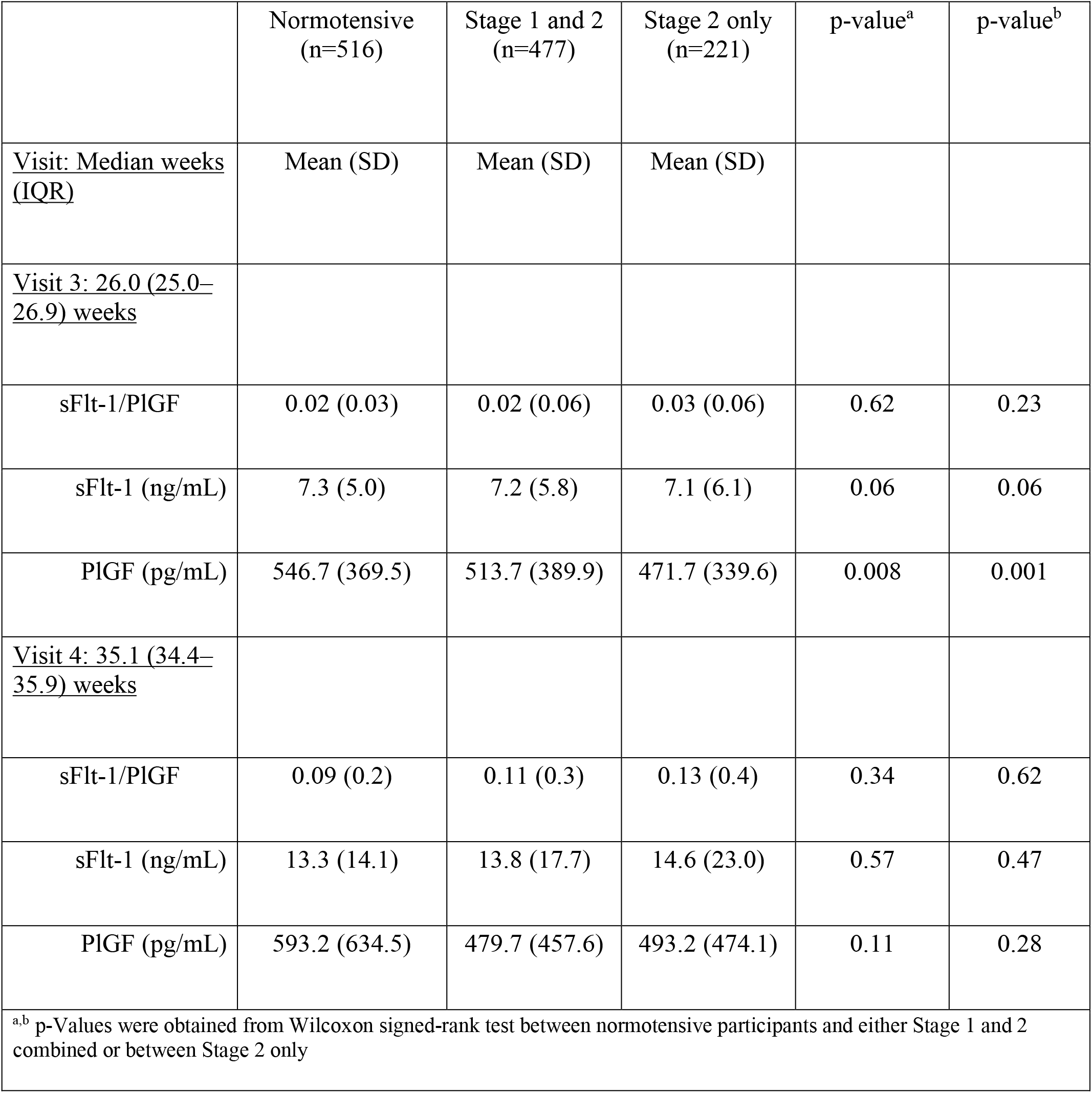
Maternal plasma sFlt-1/PlGF ratio and concentrations of sFlt-1 and PlGF in the second and third trimester by HTN status within 15 years of index pregnancy among all participants in LIFECODES, 2006–2008 (N=993)

|  | Normotensive<br>(n=516) | Stage 1 and 2<br>(n=477) | Stage 2 only<br>(n=221) | p-value<sup>a</sup> | p-value<sup>b</sup> |
| --- | --- | --- | --- | --- | --- |
| <u>Visit: Median weeks (IQR)</u> | Mean (SD) | Mean (SD) | Mean (SD) |  |  |
| <u>Visit 3: 26.0 (25.0–26.9) weeks</u> |  |  |  |  |  |
| sFlt-1/PlGF | 0.02 (0.03) | 0.02 (0.06) | 0.03 (0.06) | 0.62 | 0.23 |
| sFlt-1 (ng/mL) | 7.3 (5.0) | 7.2 (5.8) | 7.1 (6.1) | 0.06 | 0.06 |
| PlGF (pg/mL) | 546.7 (369.5) | 513.7 (389.9) | 471.7 (339.6) | 0.008 | 0.001 |
| <u>Visit 4: 35.1 (34.4–35.9) weeks</u> |  |  |  |  |  |
| sFlt-1/PlGF | 0.09 (0.2) | 0.11 (0.3) | 0.13 (0.4) | 0.34 | 0.62 |
| sFlt-1 (ng/mL) | 13.3 (14.1) | 13.8 (17.7) | 14.6 (23.0) | 0.57 | 0.47 |
| PlGF (pg/mL) | 593.2 (634.5) | 479.7 (457.6) | 493.2 (474.1) | 0.11 | 0.28 |
| <sup>a,b</sup> p-Values were obtained from Wilcoxon signed-rank test between normotensive participants and either Stage 1 and 2 combined or between Stage 2 only |  |  |  |  |  |

Table 5 demonstrates sFlt-1/PlGF ratios and concentrations by interquartile (IQR) increase and the hazard of developing HTN within 15 years of delivery. An IQR increase in sFlt-1, PlGF, or sFlt-1/PlGF ratio was not associated with an increased or decreased hazard of later life HTN. Similarly, after the models were adjusted for covariates, including maternal age at initial visit, pre-pregnancy BMI, systolic BP at initial visit, race/ethnicity, insurance status, smoking status, and diagnosis of PE/GHTN, there still was no association between an IQR increase in biomarkers and hazard of HTN.

**Table 5.**
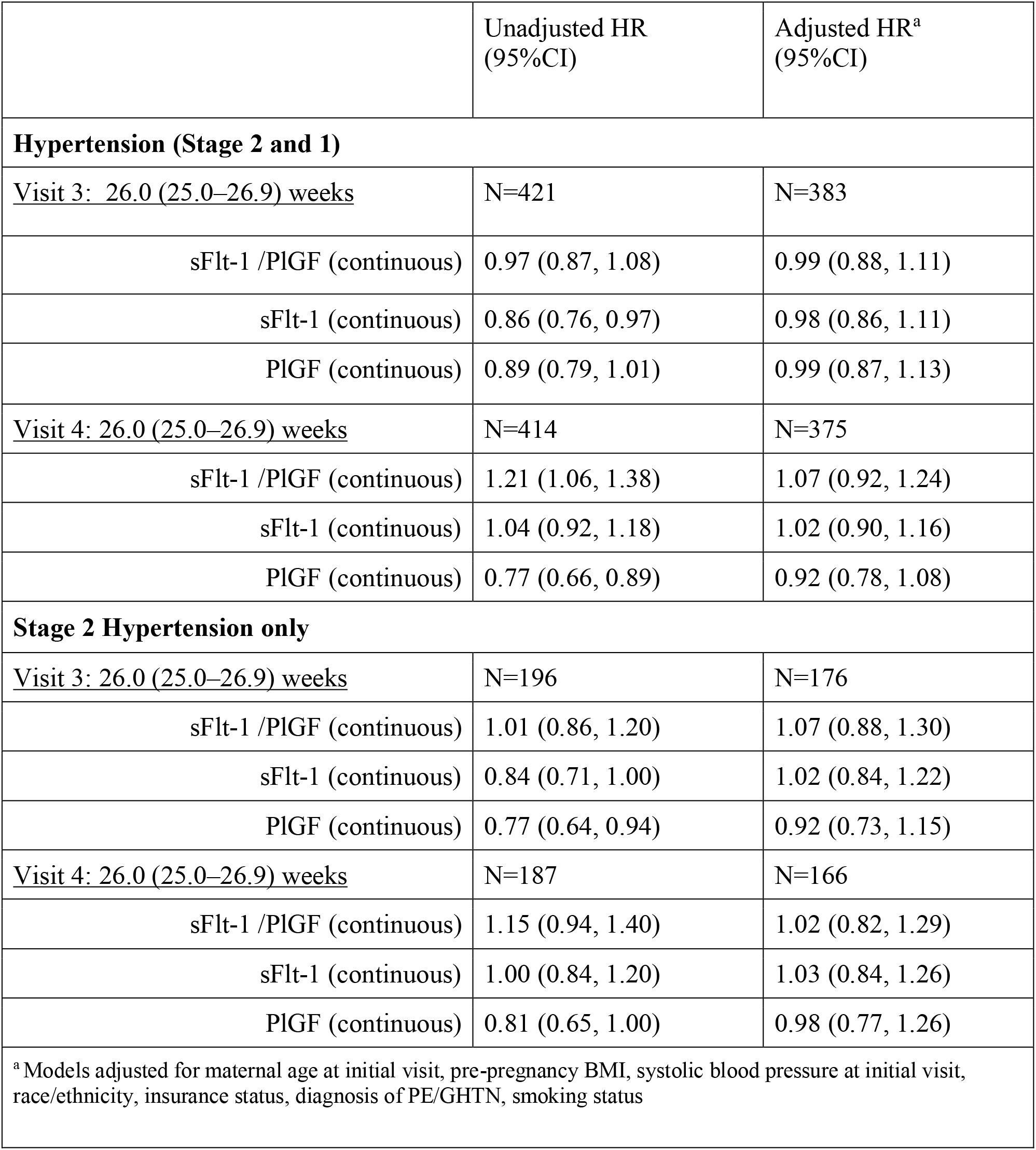
Hazard of HTN within 15 years after delivery in association with interquartile increase in angiogenic factors during pregnancy

## DISCUSSION

This study investigated the independent and comparative associations of HDP and circulating biomarkers with the risk of developing HTN in a cohort of 993 women followed for up to 15 years after delivery. Using Cox proportional hazards regression, we estimated hazard ratios for stage 1 and stage 2 HTN, adjusting for key covariates. Our findings indicate that all PE (adjusted HR=2.17; 95% CI: 1.44 -3.28) and GHTN (adjusted HR=3.50; 95% CI:2.21 -5.54) were associated with increased HTN risk. In contrast, interquartile increases in sFlt-1/PlGF ratios in the second and third trimester, regardless of the diagnosis of HDP in pregnancy, were not significantly associated with the hazard of developing HTN later in life in patients followed for up to 15 years after delivery. However, PlGF levels measured in the second and third trimester were significantly lower in those who developed HTN later in life.

PE is an established risk factor for major cardiovascular and cerebrovascular events (MACCE). In the short-term, patients with PE have an elevated risk of developing HTN, peripartum cardiomyopathy, and cardiac dysfunction in the early postpartum period^28–30^. Specifically, high levels of sFlt-1 measured in the third trimester have been shown to be associated with cardiac dysfunction and immediate PP HTN^31^. Although sFlt-1 levels fall after delivery of the placenta, altered angiogenic serum levels persisted in a follow-up study 2 years PP^32^.

These observations are like those made in mouse models. Induction of PE by over-expressing sFlt-1 in mice led to long term changes in the plasma proteome that correlate with proteome profiles observed in CVD. The mice who had experimental PE demonstrated an increased vascular proliferative and fibrotic response to unilateral carotid injury compared to normal controls^33^. Similarly, others observed that sFlt-1 induced vascular injury during pregnancy led to enhanced sensitivity of smooth muscle cell mineralocorticoid receptors, contributing to PP HTN in response to stressors such as increased salt load^34^. These studies suggest that high levels of sFlt-1 during pregnancy in mice alters the cardiovascular system after the initial PE insult.

Our biomarker observations are in concordance with prior observational work. In a secondary analysis of a prospective cohort study of 80 women, there was no difference in sFlt-1 or sFlt-1/PlGF ratio collected at a median GA of 30 weeks between those who did and did not have HTN 1 year PP. However, mean PlGF levels were lower in the 23 women who developed HTN 1 year postpartum compared to those who did not develop HTN (23 vs 48 pg/mL, p=0.017)^35^. A prospective study of 40 women found that elevated serum sFlt-1/PlGF at the time of diagnosis of PE was correlated with the mean diastolic blood pressure checked at a follow up of 12 years postpartum, which is the study with the longest follow-up of BPs PP, though with a smaller cohort^21^. It is possible that our study did not show a relationship between serum sFlt-1/PlGF ratios because samples were collected at pre-determined timepoints in the pregnancy (second and third trimester) and not necessarily proximal to or at the time of HDP diagnosis.

The current study is strengthened by its application of data from one of the largest pregnancy biobanks available, with follow up BPs for up to 15 years after delivery. Previous studies examining the development of HTN later in life used the outdated criteria for HTN, while this study incorporated the updated definition for HTN (SBP ≥130 mm Hg and/or DBP ≥ 80 mm Hg). Our study uniquely evaluated biomarkers on all patients enrolled in the database, irrespective of a diagnosis of HDP. While sFlt-1/PlGF in pregnancy was not associated with HTN later in life, our study found that PlGF was decreased in those who developed HTN later in life, regardless of concurrent diagnosis of HDP. Patients who remained normotensive in the study follow up period had significantly higher levels of PlGF in the second trimester, compared to those who went on to develop stage 1 and stage 2 HTN later in life. Thus, low PlGF may be a window into a patient’s overall vascular health and detecting lower PlGF levels in pregnancy may serve as an early harbinger of later life CVD, as we saw in this study.

The study is limited by the number of patients who had PE, particularly those with early-onset PE, which is strongly associated with later life cardiovascular disease. This may explain why we did not see a significant association between early-onset PE and later life HTN. Even more, this may have affected the ability to detect a significant association between sFlt-1/PlGF ratios and later life HTN. The retrospective design limited the amount of BP values available for follow-up. Of the 1238 patients that were enrolled in the database, we were able to retrieve follow-up BP values for 993 of the participants. The rest of the participants did not have follow-up care within the medical system and thus no follow-up data was available. The demographic data of the group with follow-up BPs was not significantly different from the demographic data of the group that was excluded from the analysis due to missing BPs or biomarker data. Once it was determined that these groups were similar, we were able to carry out the analysis of our participants with follow up data with additional understanding that the group without follow up was not more likely to be of a certain age, race, educational background, or have different comorbidities, addressing ascertainment bias. Of note, the platform used in our study to measure the serum sFlt-1/PlGF ratios was an investigational immunoassay on the Architect ci8200 platform in a research core laboratory (Abbott Laboratories, Abbott Park, IL) ^24^. This was the platform available at the time of the study and is different than the platform used in the PRAECIS study that established the serum sFlt-1/PlGF ratio cut-off of 40, which predicts the progression to severe PE within the next two weeks^20^.

The results of this study highlight the importance of comprehensive and age-appropriate screening for PP women, as no markers have reliably correlated with future risk of developing HTN. The AHA and ACOG released a joint statement recognizing pregnancy as an important milestone that shapes cardiovascular health^36^. However, current CVD risk calculators are not useful for young women after HDP for risk stratification for HTN and CVD given that these do not take into account pregnancy complications and often are not applicable to patients below the age of 40^37^. According to the “2019 ACC/AHA Guideline on the Primary Prevention of Cardiovascular Disease: A Report of the American College of Cardiology/American Heart Association Task Force on Clinical Practice Guidelines,” adults who are 40 to 75 years of age and being evaluated for CVD prevention should undergo 10-year atherosclerotic cardiovascular disease (ASCVD) risk estimation and have a clinician–patient risk discussion before starting on pharmacological therapy, such as antihypertensive therapy, a statin, or aspirin^38^. In addition, assessing for other risk-enhancing factors can help guide decisions about preventive interventions in select individuals. A history of PE as a risk factor for CVD is captured only qualitatively in the current cardiology guidelines regarding primary prevention. Per the algorithm, a history of PE is considered a “risk enhancer,” which if present, encourages shared decision making regarding statin initiation in “borderline risk” patients who otherwise would not qualify for a statin. While we aimed to see if biomarkers measured in pregnancy would enable more granular risk stratification as opposed to “risk enhancement” by clinical diagnosis, our study did not show a correlation between biomarkers and later life HTN.

Highlighted above is our poor ability to predict future risk and determine commonalities between preeclampsia and the various future CV conditions it brings with it. While clinical diagnoses of HDP were associated with HTN later in life, sFlt-1/PlGF ratios did not retain their utility in an association with future HTN. Further studies should continue to probe at these relationships using these and other biological markers. Finding a common biological marker would deepen our understanding of the interplay between PE and CVD and improve screening and the ability to offer novel targets for the prevention of CVD in patients with a history of PE.

## Data Availability

All data produced in the present study are available upon reasonable request to the authors

## Acknowledgements

None

---

**Supplement 1.**
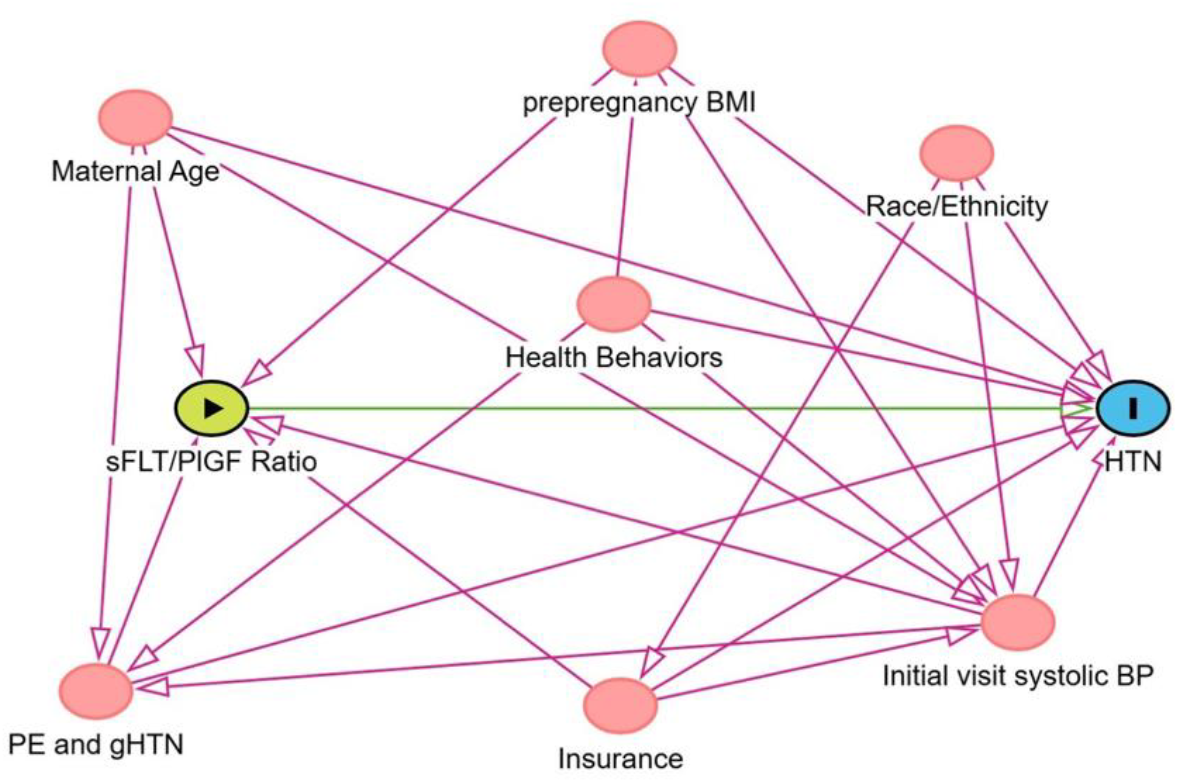
Directed Acyclic Graph BP= blood pressure, BMI= Body Mass index, PE= preeclampsia, GHTN= Gestational hypertension, sFlt-1/PLGF ratio, sFlt-1/PlGF= soluble fms-like tyrosine kinase-1 to placental growth fact

